# Inter-Rater Agreement for the Annotation of Neurologic Concepts in Electronic Health Records

**DOI:** 10.1101/2022.11.16.22282384

**Authors:** Chelsea Oommen, Quentin Howlett-Prieto, Michael D. Carrithers, Daniel B. Hier

## Abstract

The extraction of patient signs and symptoms recorded as free text in electronic health records is critical for precision medicine. Once extracted, signs and symptoms can be made computable by mapping to clinical concepts in an ontology. Extracting clinical concepts from free text is tedious and time-consuming. Prior studies have suggested that inter-rater agreement for clinical concept extraction is low. We have examined inter-rater agreement for annotating neurologic concepts in clinical notes from electronic health records. After training on the annotation process, the annotation tool, and the supporting neuro-ontology, three raters annotated 15 clinical notes in three rounds. Inter-rater agreement between the three annotators was high for text span and category label. A machine annotator based on a convolutional neural network had a high level of agreement with the human annotators, but one that was lower than human inter-rater agreement. We conclude that high levels of agreement between human annotators are possible with appropriate training and annotation tools. Furthermore, more training examples combined with improvements in neural networks and natural language processing should make machine annotators capable of high throughput automated clinical concept extraction with high levels of agreement with human annotators.

## INTRODUCTION

Extracting medical concepts from electronic health records is key to precision medicine [15]. The signs and symptoms of patients (part of the patient phenotype) are generally recorded as free text in progress notes, admission notes, and discharge summaries [22]. Clinical phenotyping of patients involves the conversion of free text into clinical concepts from an ontology [3, 31]. This is a two-step process that involves identifying appropriate text spans in narratives and then mapping the text spans to target concepts in an ontology [1, 12].

patient movements were **ataxic** ⇒ **ataxia** ⇒ UMLS CUI: **C0004134** free text ⇒ clinical concept ⇒ machine readable code

In this example, an annotator highlights the term ataxic, then maps it to the concept ataxia, and the UMLS CUI C0004134 [6]. This is a slow and error-prone process for human annotators. Agreement between human raters for annotation in clinical text is often low. A study conducted on the agreement for SNOMED CT codes between coders from three professional coding companies yielded about 50 percent agreement for exact matches with slightly higher agreement when adjusted for near matches [4]. Another study of SNOMED CT coding of ophthalmology notes yielded low levels of inter-rater agreement ranging from 33 to 64 percent [20]. Identified sources of disagreement between coders included human errors (lack of applicable medical knowledge, lack of recognition of abbreviations for concepts, and general carelessness), annotation guideline flaws (under specified and unclear guidelines), ontology flaws (polysemy of coded concepts), interface term issues (inconsistent categorization of clinical jargon), and language issues (interpretation difficulties due to use of ellipsis, anaphora, paraphrasing, and other linguistic concepts) [24].

The goal of high throughput phenotyping is to use natural language processing (NLP) to automate the annotation process [19]. Approaches to high throughput clinical concept extraction have included rule-based systems, traditional machine learning algorithms, deep learning algorithms, and hybrid methods that combine algorithms [12]. Rule-based systems such as cTAKES and MetaMap generally have accuracy and recall between 0.38 and 0.66 [1, 10, 17]. Neural networks are being used for concept recognition with increasing success. Arbabi et al. developed a convolutional neural network that matches input phrases to concepts in the Human Phenotype Ontology with high accuracy [5]. Other deep learning approaches, including neural networks based on bidirectional encoder representations from transformers (BERT), show promise for automated clinical concept extraction [12, 1, 33]. In this paper, we examine inter-rater agreement for text-span identification of neurological concepts in notes from electronic health records. In addition to the agreement between human annotators, we examine the agreement between human annotators and a machine annotator based on a convolutional neural network.

## METHODS

### Annotation tool

Prodigy (Explosion AI, Berlin, Germany) was used to annotate neurologic concepts in the EHR physician notes. Prodigy runs under python in the terminal mode of macOS, Windows, or Linux. It creates a web interface locally (Figures 1a and 1b). Annotations are stored in an SQLite database and are exportable as JSONL files. Prodigy integrates with the *spaCy* natural language processing toolkit (Explosion AI) and can train neural networks for named entity recognition and text classification.

**Figure 1a.**
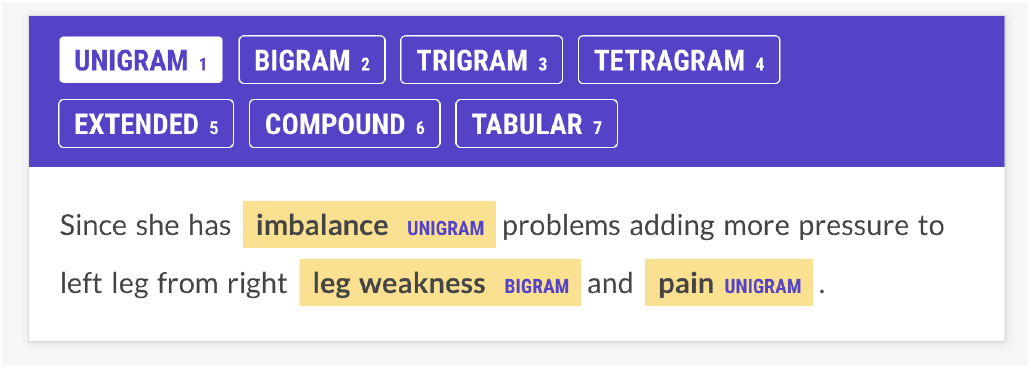
Annotator screen for a patient with multiple sclerosis. The patient complains of imbalance, leg weakness, and pain, and these concepts have been annotated. Imbalance and pain are labeled as unigrams; leg weakness is labeled a bigram. Annotators were trained to ignore laterality (e.g., right leg weakness.)

**Figure 1b.**
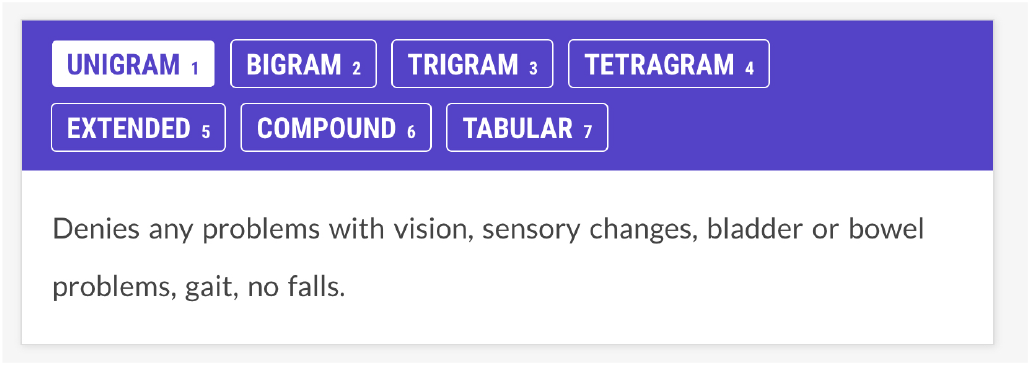
Annotator screen for neurological concepts for a patient with multiple sclerosis. The patient denies problems with vision, sensation, bladder, bowel, gait, or falls. The annotators are trained not to annotate negated concepts. The NN had no specific negation rule but learned not to tag negated concepts through training examples.

The kappa statistic was used to assess agreement between the three annotators and the neural network. The kappa statistic corrects observed rater agreement for chance rater agreement. It ranges from −1 to 1, where 1 is complete agreement, 0 is a chance agreement, and −1 indicates less-than-chance agreement. Values of kappa of 0.6 to 0.79 are considered substantial agreement, values between 0.8 and 0.90 are considered strong agreement, and values over 0.90 are considered near perfect agreement [23, 8].

### Rater training and instructions

Three annotators participated in the research. Annotator 1 (A1) was a senior neurologist, Annotator 2 (A2) was a pre-medical student majoring in neuroscience, and Annotator 3 (A3) was a third-year medical student. Raters first reviewed concepts in the neuro-ontology of neurological concepts [16] and then were instructed to find all neurological concepts in the neurology notes. Signs and symptoms (ataxia, fatigue, weakness, memory loss, etc.) were annotated but not disease entities (Alzheimer’s disease, multiple sclerosis, etc.) Raters annotated the neurologic concepts and ignored laterality and other modifiers (e.g., *arm pain* for *right arm pain, back pain* for *severe back pain*, etc.) In addition, annotators tagged each text span with an category label (see Figure 1a and 1b). Category labels included *unigrams* (one-word concepts such as ataxia), *bigrams* (two-word concepts such as double vision), *trigrams* (three-word concepts such as low back pain), *tetragrams* (four-word concepts such as relative afferent pupil defect), *extended* (text span annotations longer than four words), *compound* (multiple concepts in one text span such as brisk ankle and knee reflex), and *tabular* (concepts represented in tabular or columnar format, usually showed right and left body sides).

### The machine annotator

The machine annotator (NN) was a neural network that was trained to recognize text spans containing neurology concepts in the electronic health record physician notes. The NN was the default spaCy named entity recognition model based on a four-layer convolutional neural network (CNN) that looked at four words on either side of each token using *tok2vec* with an initial learning rate 1 ∗ *e*^−3^. NN was trained on 11,000 manually annotated sentences derived from neurology textbooks, online neurological disease descriptions, and electronic health record notes.

### Annotations

Five patient EHR notes were annotated for each of the three rounds. The annotation of EHR clinical notes for research purposes was approved by the Institutional Review Board of the University of Illinois. Informed patient consent for use of clinical notes was obtained from all subjects through the UIC Biobank Project. Three human annotators (A1, A2, and A3) and the machine annotator (NN) annotated each note. After each round, the annotators met and reviewed any annotation disagreements. EHR notes varied in length. The number of concepts to annotate for Round 1 was 117, for Round 2 was 129, and for Round 3 was 114. The annotations of each annotator were stored in an SQLite database and exported as a JSON file for scoring for inter-rater agreement in python. Text spans were mapped to concepts in the neuro-ontology [16] utilizing a lookup table with 3,500 target phrases and the similarity method from spaCy [2] (pp. 152-154). Univariate analysis of variance and Cohen’s kappa statistics were calculated with SPSS (IBM, version 28).

## RESULTS

Annotators identified neurological concepts in physician notes from electronic health records. Each annotator identified the text span associated with the neurological concept and assigned a category label to each annotation (e.g., unigram, bigram, trigram, etc.) Inter-rater agreement was calculated between the three human annotators and between the machine annotator (NN).

Unadjusted agreement on the text span task was 88.9% ± 3.2 between the human annotators and was 83.9% ± 4.6 between the human annotators and the machine annotator (human-human mean was higher, one-way ANOVA, df=1, p = 0.016). Unadjusted agreement on the category label task was 87.7% ± 4.4 between human annotators and was 84.6% ± 5.5 between the human annotators and the machine annotator (means did not differ, one-way ANOVA, df=1, p= 0.212).

Cohen’s kappa statistic (*κ*) was high for both the text span task (0.715 to 0.893) and the category label task (0.72 to 0.89) (Figures 2a and 2b). On the text span identification task (Figure 3a) *κ* was higher for the human-human pairs (0.85 ± 0.05) than the human-machine pairs (0.76 ± 0.06). On the category label task, *κ* (Figure 3b) was similar between the human-human pairs (0.83 ± 0.05) and the human-machine pairs (0.82 ± 0.06). *κ* for the text span task (Figure 4a) or the category label task (Figure 4b did not differ by round (one way ANOVA, df=2, p *>*0.05).

**Figure 2a.**
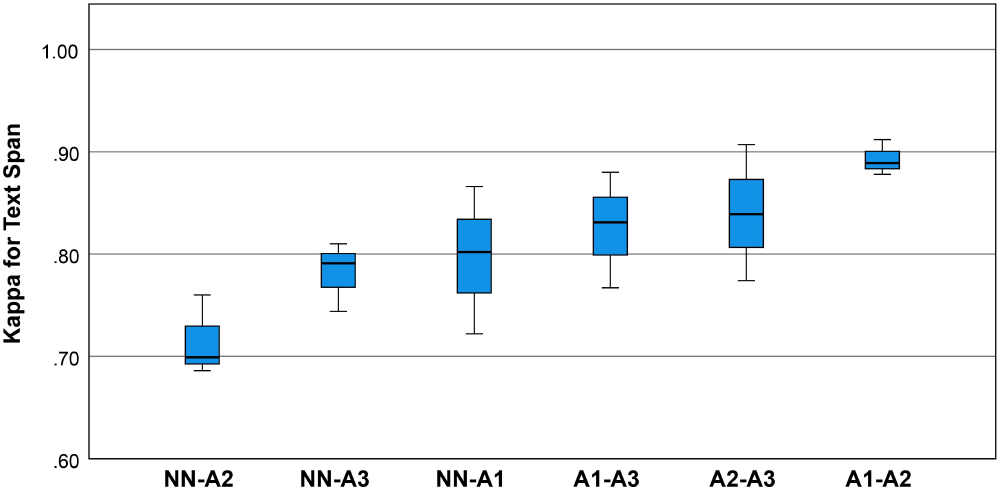
Boxplots for the kappa statistic for inter-rater agreement for text spans for the neurological concepts. Univariate analysis of variance showed that mean inter-rater agreement differed by rating pair (one-way ANOVA, df=5, p = 0.021). Post hoc comparisons by the Bonferroni method showed that pair A1-A2 outperformed pair NN-A2.

**Figure 2b.**
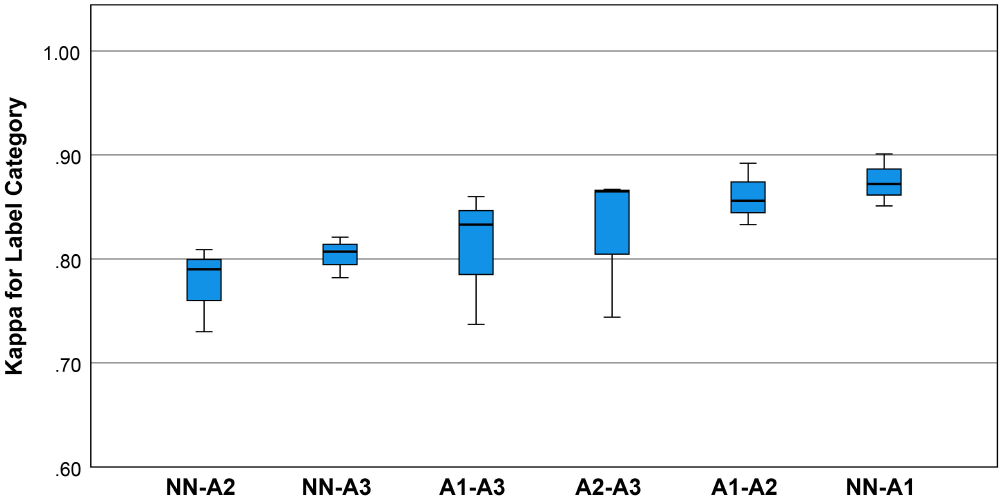
Boxplots for the kappa statistic for inter-rater agreement for category labels for the neurological concepts. Univariate analysis of variance showed that mean kappa for category label agreement did not differ by rating pair (one-way ANOVA, p = 0.165, df=5).

## DISCUSSION

Signs and symptoms are an important component of a patient’s phenotype. Extracting these phenotypic features from electronic health records and converting them to machine-readable codes makes them computable [18]. These computable phenotypes are critical to precision medicine initiatives [14, 28, 9]. Agrawal et al. [1] have conceptualized clinical entity extraction as a two-step process of text span recognition followed by clinical entity normalization. Text span recognition is the identification of clinical concepts in the free text; entity normalization is the mapping of this text to clinical concepts in an ontology such as UMLS [6]. We have focused on an inter-rater agreement for text span annotation. For entity normalization, we depended on a look-up table that mapped text spans to concepts in neuro-ontology. We found high inter-rater agreement among the human annotators (approximately 89%) with a lower agreement between the human annotators and the machine annotator (approximately 84%). The kappa statistic for human-human rater agreement was between 0.77 and 0.91, and the kappa statistic for the human-machine agreement was between 0.69 and 0.87 (Figure 3a). We consider the inter-rater agreement between the human raters (0.77 to 0.91) as good, especially when contrasted with the inter-rater agreement between trained neurologists eliciting patient signs and symptoms [30, 13]. For trained neurologists eliciting signs and symptoms such as weakness, sensory loss, ataxia, aphasia, dysarthria, and drowsiness, the *κ* statistics range from 0.40 to 0.70 [30, 13].

**Figure 3a.**
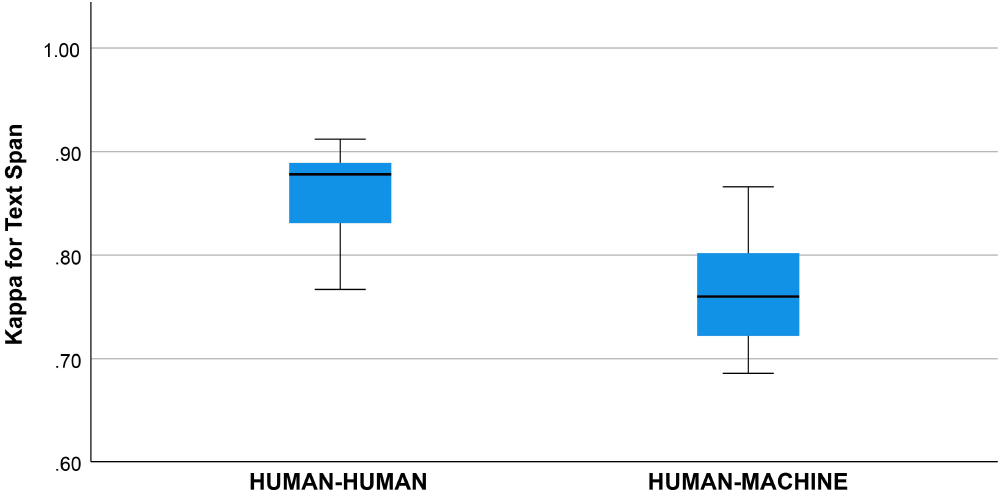
Kappa statistic for agreement between human-human and human-machine raters for text span. Groups differed, one-way ANOVA, df=1, p = 0.004.

**Figure 3b.**
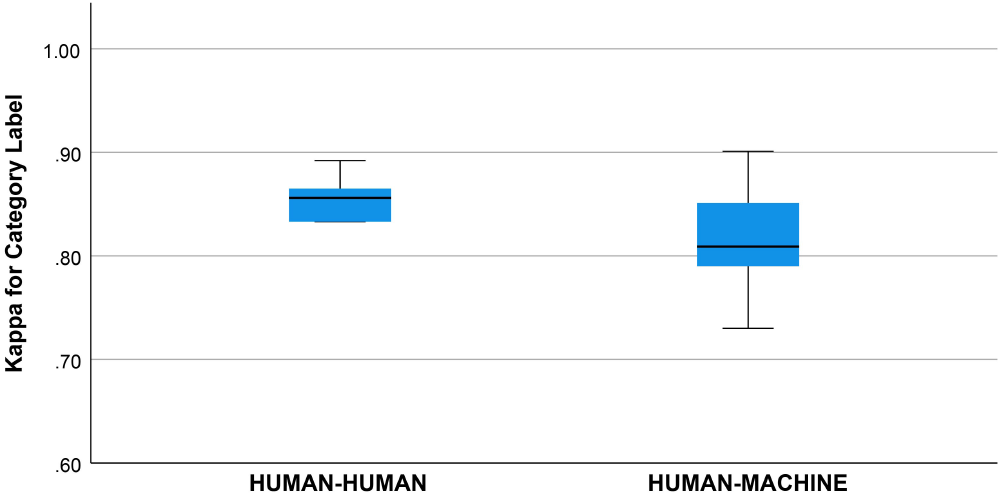
Kappa statistic for agreement between human-human and human-machine raters for category label. Groups did not differ, one way ANOVA, df=1, p = 0.589

We did not find a training effect for the human annotators across rounds (Figure 4a and 4b). Although the annotators met after each round and discussed discrepancies in their annotations, inter-rater agreement did not improve significantly between rounds. This suggests that there may be a ceiling for inter-rater agreement for text span annotation with a kappa of 0.80 to 0.90 and that higher levels of agreement may not be possible due to the difficulty of the task and random factors that cannot be addressed by additional training or experience. This ceiling effect for the human inter-rater agreement has implications for the potential for higher rates of inter-rater agreement between humans and machines (Figure 3b). Mean inter-rater agreement for text span was higher for the human-human pairs (*κ* = 0.85) than the human-machine pairs (*κ* = 0.76). Additional training examples are likely to improve the performance of the machine annotator on the text span and category label tasks. Furthermore, other neural networks may outperform the convolutional neural network (CNN), which is the baseline for Prodigy. We have preliminary information that a neural network based on bidirectional encoder representations from transformers (BERT) can improve performance on the text span task by 5 to 10% (unpublished results). Others have found that deep learning approaches based on BERT outperform approaches based on CNN for concept identification and extraction tasks [33]. We think a ceiling effect for inter-rater agreement, whether human-human or human-machine, near a *κ* of 0.90 is likely.

**Figure 4a.**
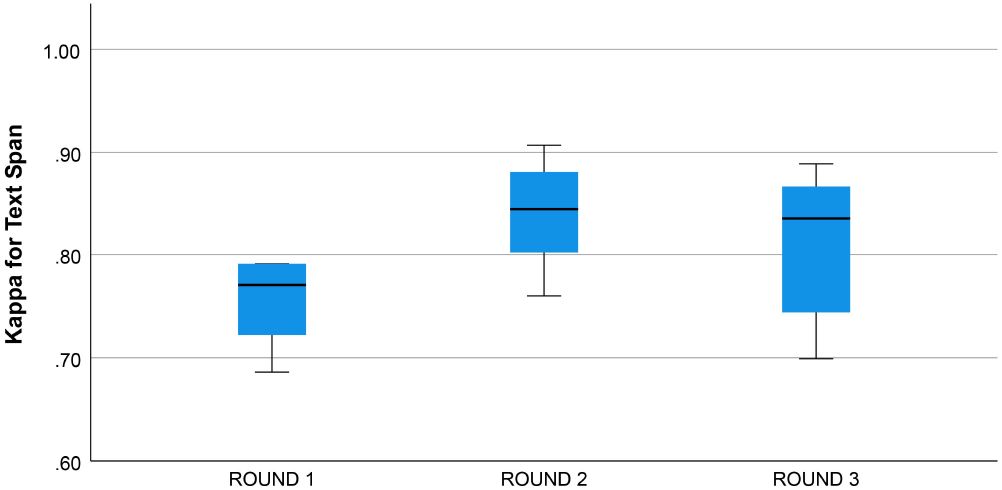
4a. Kappa statistic for inter-rater agreement for text span by round. Groups do not differ, one-way ANOVA, df=2, p = 0.30.

**Figure 4b.**
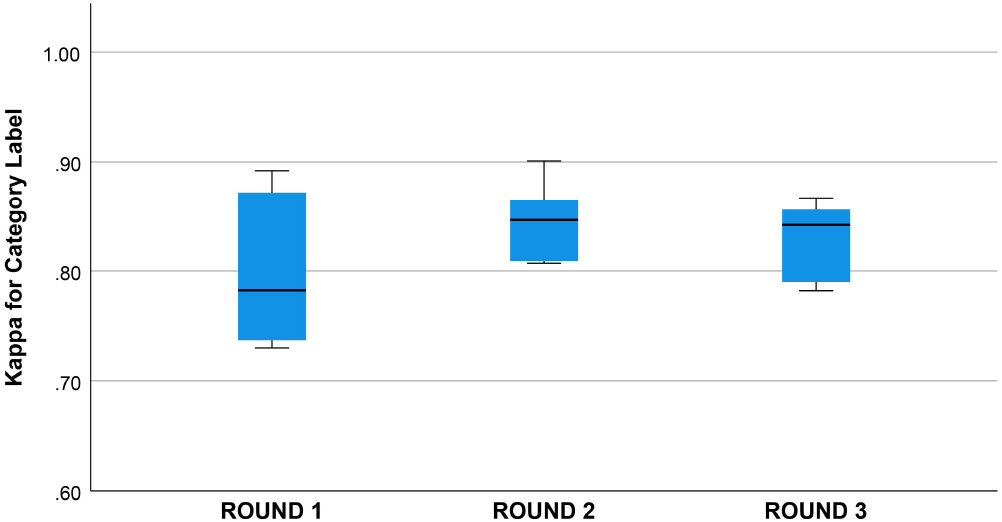
Kappa statistic for inter-rater agreement for category label by round. Groups do not differ, one-way ANOVA, df=2, p = 0.31.

Given the heavy documentation burden on physicians and physician burn-out attributed to electronic health records, physician documentation of signs and symptoms is likely to continue as free text. Structured documentation of signs and symptoms as an alternative to free text is too burdensome in the current environment [32, 7, 21, 29, 25, 11]. A medium-sized medical center with a daily inpatient census of 300 and a daily outpatient census of 2,000 generates at least 5,000 clinical notes daily or over 1.5 million notes annually (unpublished estimates based on two academic medical centers). The sheer volume of clinical notes in electronic health records makes the manual annotation of clinical concepts impractical. Extracting signs and symptoms for precision medicine initiatives will depend on advances in natural language processing and natural language understanding.

Although high throughput phenotyping of electronic health records by manual methods is impractical, [19], the manual annotation of free text in electronic health records can be used to train neural networks for phenotyping. Neural networks can also speed up the manual annotation process. The annotator Prodigy [26, 27] has an annotation mode called *ner*.*correct*, which uses a trained neural network to accelerate the manual annotation of clinical concepts.

With suitable training and guidelines, high levels of inter-rater agreement between human annotators for clinical concepts are feasible. Restricting the annotation to a limited domain (e.g., neurological concepts) and restricted ontology (e.g., neuro-ontology) simplifies the manual annotation process. Although the inter-rater agreement between human and machine annotators was lower than between human annotators, advances in natural language processing should bring inter-rater agreement between machines and humans closer and make high throughput phenotyping of electronic health records feasible.

This work has limitations. The sample of clinical notes was small (five patient notes per annotation round). A larger sample of notes would have been desirable. The annotation process was restricted to neurological signs and symptoms in neurology notes. The target ontology was a limited neuro-ontology with 1600 concepts [16]. We evaluated only one machine annotator based on a convolutional neural network. Other neural networks are likely to perform better. Our results on an inter-rater agreement might not generalize to other medical domains and other ontologies. Although we had three raters for this study, we did not designate any of them as the “gold standard”, and we elected to calculate inter-rater agreement for each pair of raters separately. In our opinion, agreement at the 90% level between human raters should be considered high. Likewise, machine annotators that can reach a 90% agreement with human annotators should be considered accurate.

## Data Availability

All data produced in the present study are available upon reasonable request to the authors

## AUTHOR CONTRIBUTIONS

Concept and design by DBH. Data collection by DBH, CO, and QHP. Data analysis by CO and DBH. Data interpretation by DBH, MDC, QHP, and CO. Initial draft by DBH and CO. Revisions, re-writing, and final approval by DBH, CO, QHP, and MDC.

## DATA AVAILABILITY STATEMENT

Supporting data is available from the corresponding author upon request.

## REFERENCES

[1] Agrawal, M., O’connell, C., Fatemi, Y., Levy, A., and Sontag, D. Robust benchmarking for machine learning of clinical entity extraction. In Machine Learning for Healthcare Conference (2020), PMLR, pp. 928–949.

[2] Altinok, D. Mastering spaCy. Packt Publishing, Birmingham UK, 2021.

[3] Alzoubi, H., Alzubi, R., Ramzan, N., West, D., Al-Hadhrami, T., and Alazab, M. A review of automatic phenotyping approaches using electronic health records. Electronics 8, 11 (2019), 1235.

[4] Andrews, J. E., Richesson, R. L., and Krischer, J. Variation of snomed ct coding of clinical research concepts among coding experts. Journal of the American Medical Informatics Association 14, 4 (2007), 497–506.

[5] Arbabi, A., Adams, D. R., Fidler, S., Brudno, M., et al. Identifying clinical terms in medical text using ontology-guided machine learning. JMIR medical informatics 7, 2 (2019), e12596.

[6] Bodenreider, O. The unified medical language system (umls): integrating biomedical terminology. Nucleic Acids Research 32, suppl 1 (2004), D267–D270.

[7] Cohen, G. R., Friedman, C. P., Ryan, A. M., Richardson, C. R., and Adler-Milstein, J. Variation in physicians’ electronic health record documentation and potential patient harm from that variation. Journal of general internal medicine 34, 11 (2019), 2355–2367.

[8] Cohen, J. A coefficient of agreement for nominal scales. Educational and psychological measurement 20, 1 (1960), 37–46.

[9] Collins, F. S., and Varmus, H. A new initiative on precision medicine. New England journal of medicine 372, 9 (2015), 793–795.

[10] Divita, G., Zeng, Q. T., Gundlapalli, A. V., Duvall, S., Nebeker, J., and Samore, M. H. Sophia: a expedient umls concept extraction annotator. In AMIA Annual Symposium Proceedings (2014), vol. 2014, American Medical Informatics Association, p. 467.

[11] Downing, N. L., Bates, D. W., and Longhurst, C. A. Physician burnout in the electronic health record era: are we ignoring the real cause? Annals of Internal Medicine 169, 1 (2018), 50–51.

[12] Fu, S., Chen, D., He, H., Liu, S., Moon, S., Peterson, K. J., Shen, F., Wang, L., Wang, Y., Wen, A., et al. Clinical concept extraction: a methodology review. Journal of biomedical informatics 109 (2020), 103526.

[13] Goldstein, L. B., Bertels, C., and Davis, J. N. Interrater reliability of the NIH stroke scale. Archives of neurology 46, 6 (1989), 660–662.

[14] Haendel, M. A., Chute, C. G., and Robinson, P. N. Classification, ontology, and precision medicine. New England Journal of Medicine 379, 15 (2018), 1452–1462.

[15] Hebbring, S. J., Rastegar-Mojarad, M., Ye, Z., Mayer, J., Jacobson, C., and Lin, S. Application of clinical text data for phenome-wide association studies (PheWASs). Bioinformatics 31, 12 (2015), 1981–1987.

[16] Hier, D. B., and Brint, S. U. A neuro-ontology for the neurological examination. BMC Medical Informatics and Decision Making 20, 1 (2020), 1–9.

[17] Hier, D. B., Yelugam, R., Azizi, S., Carrithers, M. D., and Wunsch II, D. C. High throughput neurological phenotyping with metamap. European Scientific Journal 18 (2022), 37–49. Accessed August 12, 2022.

[18] Hier, D. B., Yelugam, R., Azizi, S., and Wunsch, D. C. A focused review of deep phenotyping with examples from neurology. European Scientific Journal 18, 4 (2022), 4–19.

[19] Hier, D. B., Yelugam, R., Azizi, S., and Wunsch II, D. C. A focused review of deep phenotyping with examples from neurology. European Scientific Journal 18 (2022), 4–19. Accessed August 12, 2022.

[20] Hwang, J. C., Alexander, C. Y., Casper, D. S., Starren, J., Cimino, J. J., and Chiang, M. F. Representation of ophthalmology concepts by electronic systems: intercoder agreement among physicians using controlled terminologies. Ophthalmology 113, 4 (2006), 511–519.

[21] Joukes, E., Abu-Hanna, A., Cornet, R., and De Keizer, N. F. Time spent on dedicated patient care and documentation tasks before and after the introduction of a structured and standardized electronic health record. Applied clinical informatics 9, 01 (2018), 046–053.

[22] Kimia, A. A., Savova, G., Landschaft, A., and Harper, M. B. An introduction to natural language processing: how you can get more from those electronic notes you are generating. Pediatric emergency care 31, 7 (2015), 536–541.

[23] Mchugh, M. L. Interrater reliability: the kappa statistic. Biochemia medica 22, 3 (2012), 276–282.

[24] Miñarro-Giménez, J. A., Martínez-Costa, C., Karlsson, D., Schulz, S., and Gøeg, K. R. Qualitative analysis of manual annotations of clinical text with snomed ct. Plos one 13, 12 (2018), e0209547.

[25] Moy, A. J., Schwartz, J. M., Chen, R. J., Sadri, S., Lucas, E., Cato, K. D., and Rossetti, S. C. Measurement of clinical documentation burden among physicians and nurses using electronic health records: a scoping review. Journal of the American Medical Informatics Association 28, 5 (2021), 998–1008.

[26] Musabeyezu, F. Comparative study of annotation tools and techniques. Master’s thesis, African University of Science and Technology, 2019.

[27] Neves, M., and Ševa, J. An extensive review of tools for manual annotation of documents. Briefings in bioinformatics 22, 1 (2021), 146–163.

[28] Robinson, P. N. Deep phenotyping for precision medicine. Human mutation 33, 5 (2012), 777–780.

[29] Rosenbloom, S. T., Denny, J. C., Xu, H., Lorenzi, N., Stead, W. W., and Johnson, K. B. Data from clinical notes: a perspective on the tension between structure and flexible documentation. Journal of the American Medical Informatics Association 18, 2 (2011), 181–186.

[30] Shinar, D., Gross, C. R., Mohr, J. P., Caplan, L. R., Price, T. R., Wolf, P. A., Hier, D. B., Kase, C. S., Fishman, I. G., Wolf, C. L., et al. Interobserver variability in the assessment of neurologic history and examination in the Stroke Data Bank. Archives of Neurology 42, 6 (1985), 557–565.

[31] Shivade, C., Raghavan, P., Fosler-Lussier, E., Embi, P. J., Elhadad, N., Johnson, S. B., and Lai, A. M. A review of approaches to identifying patient phenotype cohorts using electronic health records. Journal of the American Medical Informatics Association 21, 2 (2014), 221–230.

[32] Vuokko, R., MäKelä-Bengs, P., Hyppönen, H., Lindqvist, M., and Doupi, P. Impacts of structuring the electronic health record: Results of a systematic literature review from the perspective of secondary use of patient data. International journal of medical informatics 97 (2017), 293–303.

[33] Yang, X., Bian, J., Hogan, W. R., and Wu, Y. Clinical concept extraction using transformers. Journal of the American Medical Informatics Association 27, 12 (2020), 1935–1942.

